# Ebola Disease Knowledge and Vaccine Acceptance in the Democratic Republic of the Congo

**DOI:** 10.64898/2026.10.15.26365673

**Authors:** Sydney Merritt, Jason Kindrachuk, Dalau Nkamba, Nicole A. Hoff, Handdy Kalengi, Megan Halbrook, Leonard Kamba, Michael Beya, Skylar Petrichko, Hannah L Wallace, Christophe Luhata, Didine K Kaba, Anne W Rimoin

**Author notes:** **CORRESPONDING AUTHOR:** Jason Kindrachuk; Anne W. Rimoin. Indicates co-first authors who contributed equally to this work (Sydney Merritt, Jason Kindrachuk). Denotes co-last authors who jointly supervised this work (Didine K Kaba, Anne W Rimoin).

## Abstract

We report general acceptance (79.0%) of Ebola vaccination in the Democratic Republic of the Congo (DRC) between 2023-2024 (n=5,062). Acceptance varied significantly by province with acceptance ranging from 47.6% in Kongo Central to 92.1% in Tshuapa province and was highest among healthcare workers. While Ebola vaccine acceptance and disease knowledge are relatively strong nationally in the DRC, regional differences do exist and may impact future vaccine rollout.

## INTRODUCTION

The re-emergence of *Orthoebolavirus bundibugyoense* (Bundibugyo virus; BDBV) in Ituri Province, Democratic Republic of the Congo (DRC) 14 years after the first detection in DRC has resulted in the largest Ebola disease outbreak in the country’s history, and second largest across all recorded Ebola disease outbreaks. The current outbreak has also expanded at an unprecedented rate, and has since been classified as the fastest growing Ebola disease outbreak in history(1). While Bundibugyo virus disease (BVD) was historically associated with lower case fatality rates (CFR) than Ebola virus disease (EVD; etiologic agent *Orthoebolavirus zairenese*, EBOV), the current CFR for the 2026 BVD outbreak in the DRC is estimated to be 48%(2). Outbreak containment and mitigation efforts are underway, yet the outbreak has continued to expand due to many contributing factors including extensive resource limitations, lack of medical countermeasures, limited healthcare access, regional instability, and community mistrust(3).

There has been considerable interest in deploying the licensed EBOV vaccines for use in the 2026 BVD outbreak. Data from rVSVΔG-ZEBOV-GP (ERVEBO), a recombinant replication-competent viral vector vaccine licensed for prevention of EVD, has shown strong efficacy in ring vaccination trials and prior use in EVD outbreaks(4-9). However, the rVSVΔG-ZEBOV-GP vaccine is not currently approved for use against non-EBOV orthoebolaviruses, including BDBV. Recent investigations have suggested that rVSVΔG-ZEBOV-GP vaccination results in the generation of cross-reactive antibodies to BDBV in vaccine recipients(10, 11); however, further analyses are needed to assess whether cross-reactive antibodies are a correlate of immune protection. As these observations and animal challenge studies have suggested that rVSVΔG-ZEBOV-GP may provide some cross-protective activity against BDBV(12, 13), 20,000 doses of the vaccine have been allocated for a Phase 3 clinical trial in DRC along with 50,000 doses for frontline and healthcare workers(14, 15). Additional BDBV vaccine candidates have also entered Phase I clinical trials including the mRNA-1469 (Moderna) and ChAdOOx1 (Oxford Vaccine Group and Pandemic Sciences Institute) vaccine candidates(16).

Given the extensive interest in rapid vaccine deployments to help facilitate BVD outbreak containment efforts and concerns regarding community mistrust, there is a critical need to assess vaccine acceptance and uptake considerations across the diverse populations found in the DRC.

### THE STUDY

We utilized data from a larger longitudinal national telephone survey on COVID-19-related topics and general vaccine acceptance for our cross-sectional analysis. From December 2023– February 2024, trained interviewers contacted 5,226 adult participants and administered surveys in French, Swahili, Lingala, Kikongo, or Tshiluba. Healthcare workers (HCWs) were the primary target population for the study, and participants from all 26 provinces in the DRC were represented. Participants were selected from historical telephone survey records of >10,000 persons and survey questions addressed behavioral and social drivers of vaccination and attitudes toward new vaccine introductions. Only those participants with complete responses to vaccine acceptance questions were included in this complete-case analysis (n = 5,062). Ebola vaccine acceptance was measured by asking participants about their interest in inclusion of an Ebola vaccine—with no specification of which vaccine—in the national vaccination schedule.

Response choices were “yes” (including for adults, for children, or both), “no”, or “I do not know this disease”, with all “yes” responses collapsed for this analysis. Attitudes, vaccine acceptance by province, and sociodemographic characteristics were assessed by 3-level or 5-level Likert scale questions and a χ2 test to assess significance (α = 0.05), as has been previously described(17).

The median age of survey respondents was 42 years (IQR 35-51) and participants were predominantly male (79.4%) **(Table 1)**. Most respondents were from Kwilu Province (12.7%) and Kasai Oriental Province (11.6%) with similar distributions among urban (50.4%) and rural (49.6%) geographic areas. Most respondents reported some post-secondary education (41.2%), followed by those who received a high school diploma (30.4%) or those having received a Bachelor’s or Advanced degree (21.4%). More than half of the respondents identified as an HCW (55.4%) with the next closest identified occupation being a community or essential services leader (14.9%). Regarding health information, most respondents had received a COVID-19 vaccination (86.9%) and did not report being immunocompromised (89.4%).

**Table 1:**
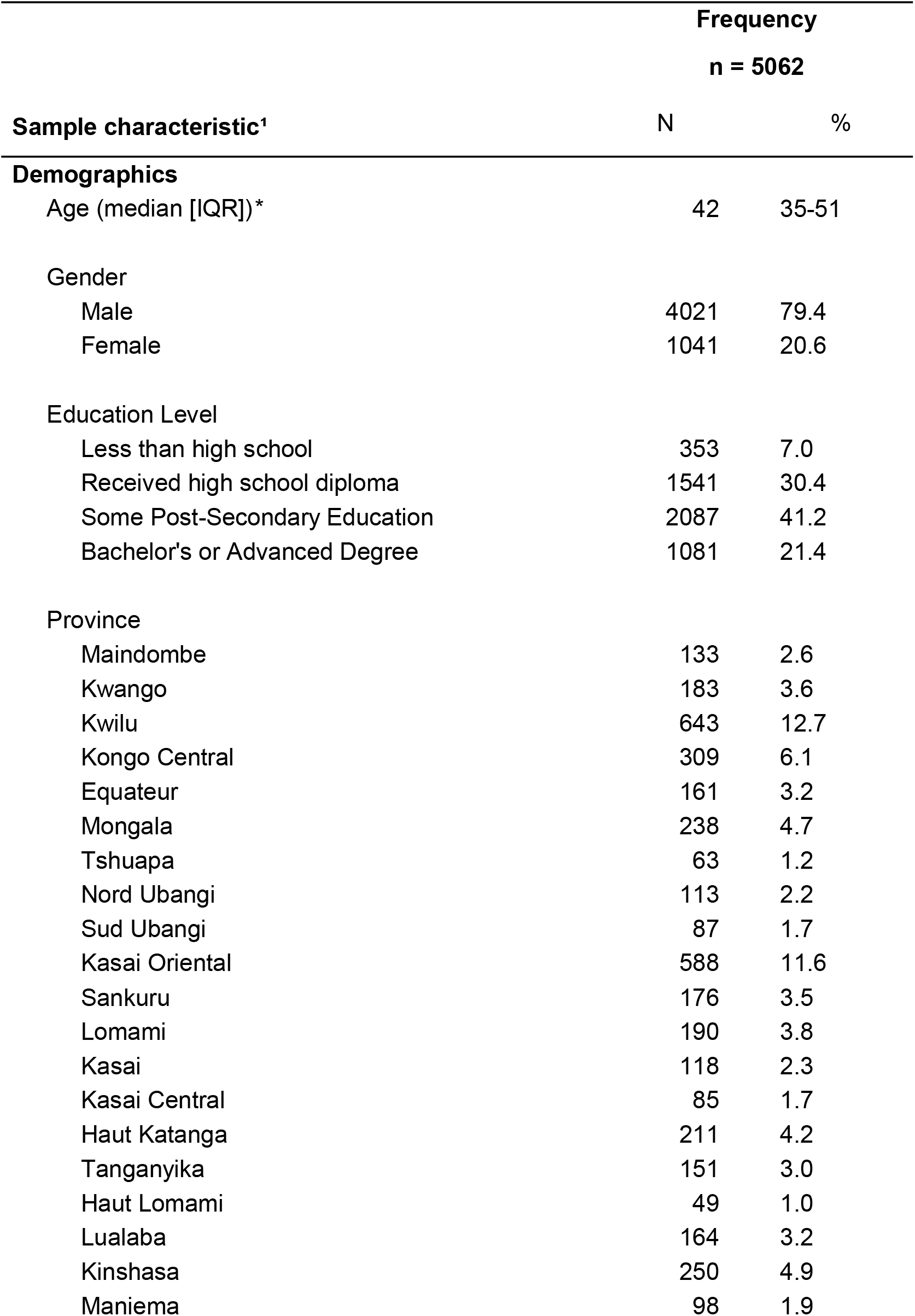
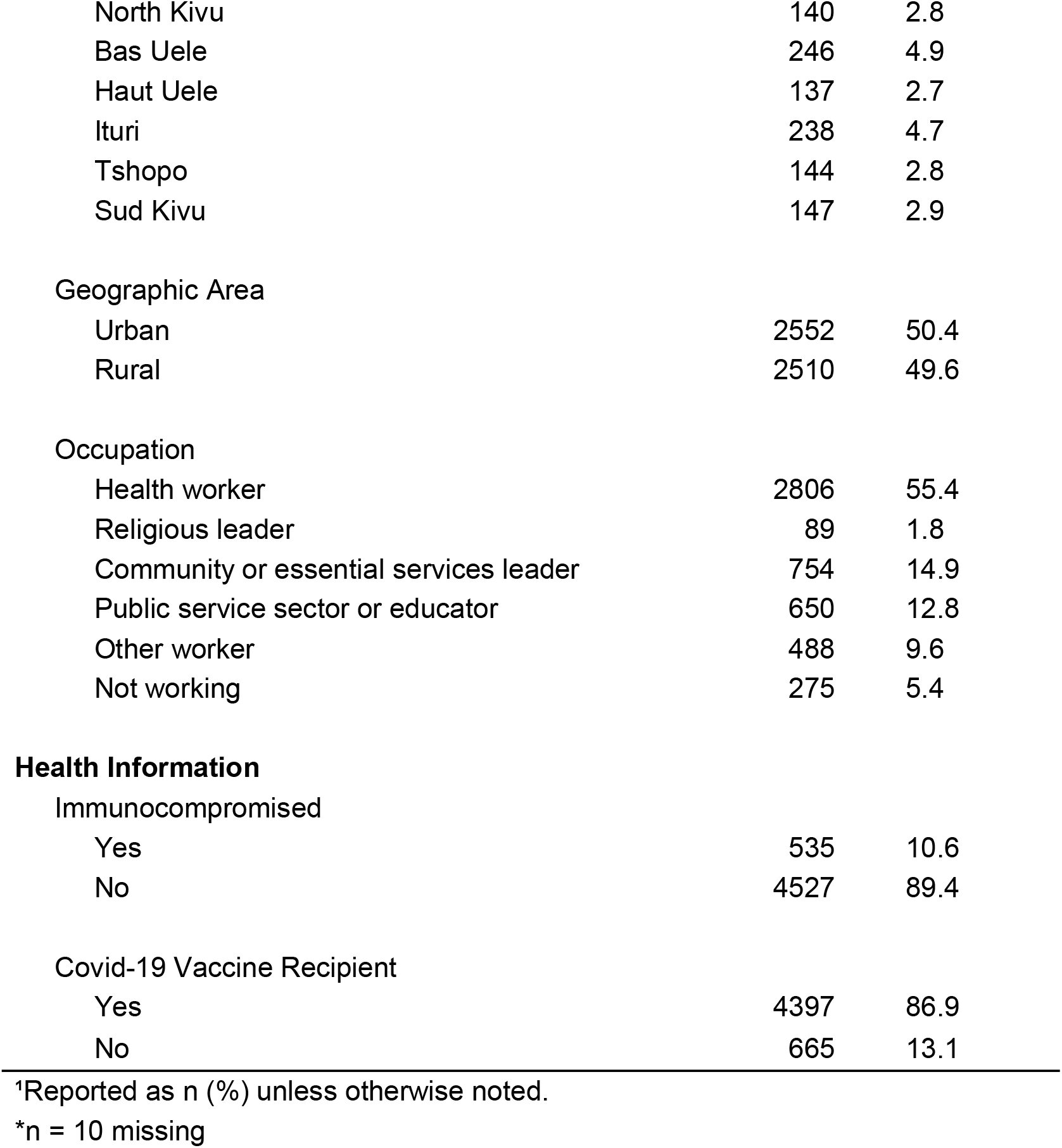
Characteristics of participants in a telephone phone survey about Ebola vaccine acceptance conducted during December 2023–February 2024, Democratic Republic of the Congo. Values are no. (%) except as indicated. IQR, interquartile range. COVID-19 vaccination was dichotomized by whether the respondent received the COVID-19 vaccine. Responses were stratified by sociodemographic characteristics and health conditions were dichotomized on the basis of whether the respondent stated they had a chronic disease or were immunocompromised.

Nationally, 3998 of respondents expressed acceptance of Ebola vaccination (79.0%), 884 had no interest, and 180 reported no Ebola knowledge. Provincially, Ebola vaccine acceptance was >90% among respondents from Tshuapa (92.1%), Haut Uele (92.0%), Mongala (90.8%), and Haut Katanga (90.5%) (Figure 1A). Two provinces had <60% Ebola vaccine acceptance - Kongo Central (47.6%) and Sud Ubangi (55.2%). Sankuru Province had the highest proportion of respondents reporting no Ebola knowledge (35.4%) (Figure 2).

**Figure 1:**
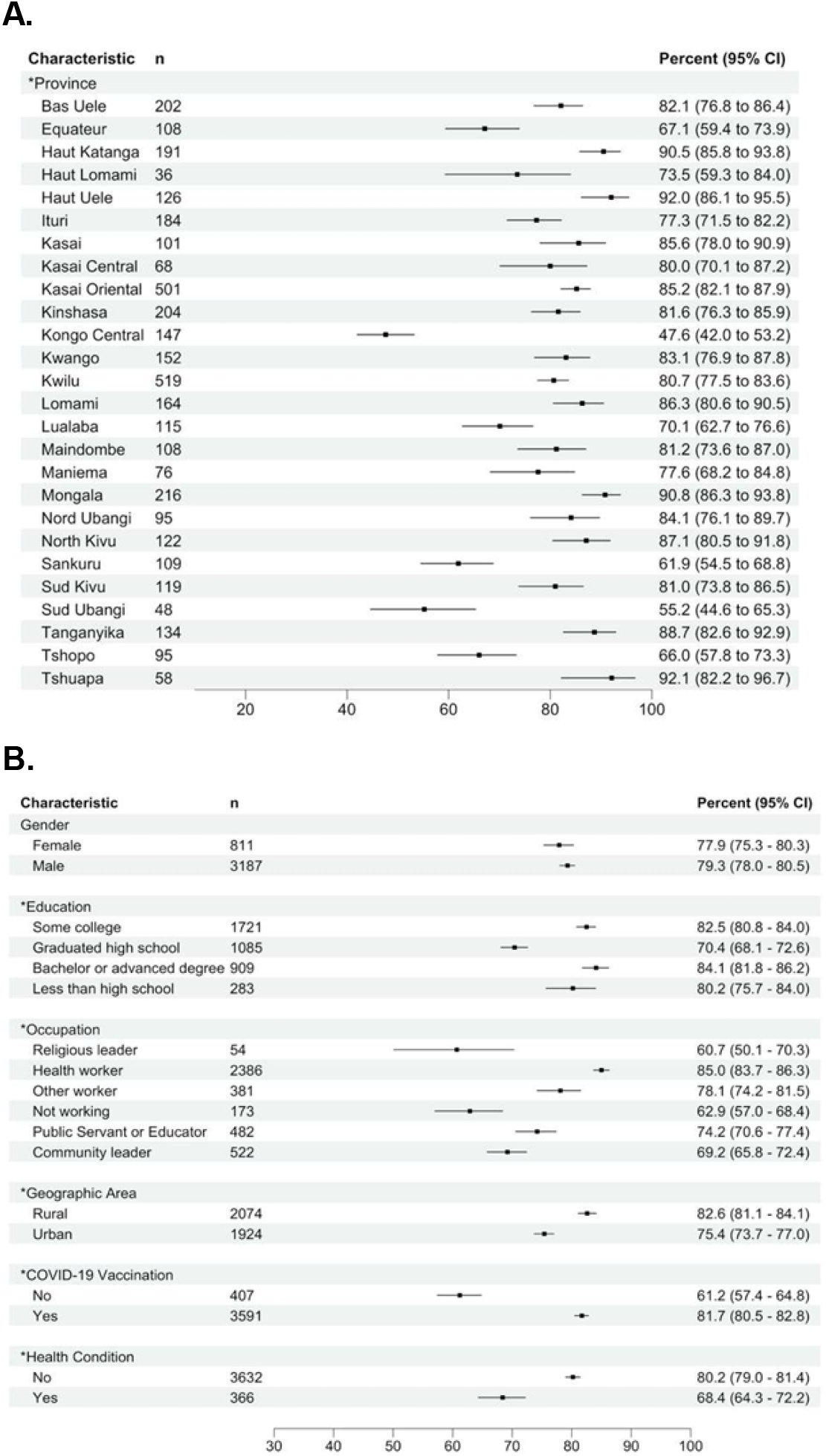
Percentage of Ebola vaccine acceptance and demographic characteristic perceptions from a telephone phone survey conducted during December 2023–February 2024, by province, the Democratic Republic of the Congo. Responses were stratified by sociodemographic characteristics and known Ebola risk factors; COVID-19 vaccination was dichotomized by whether the respondent received the COVID-19 vaccine. Health conditions were dichotomized on the basis of whether the respondent stated they had a chronic disease orwere immunocompromised. Asterisks (*) indicate p<0.05 by χ2 test. Error bars indicate 95% CIs.

**Figure 2:**
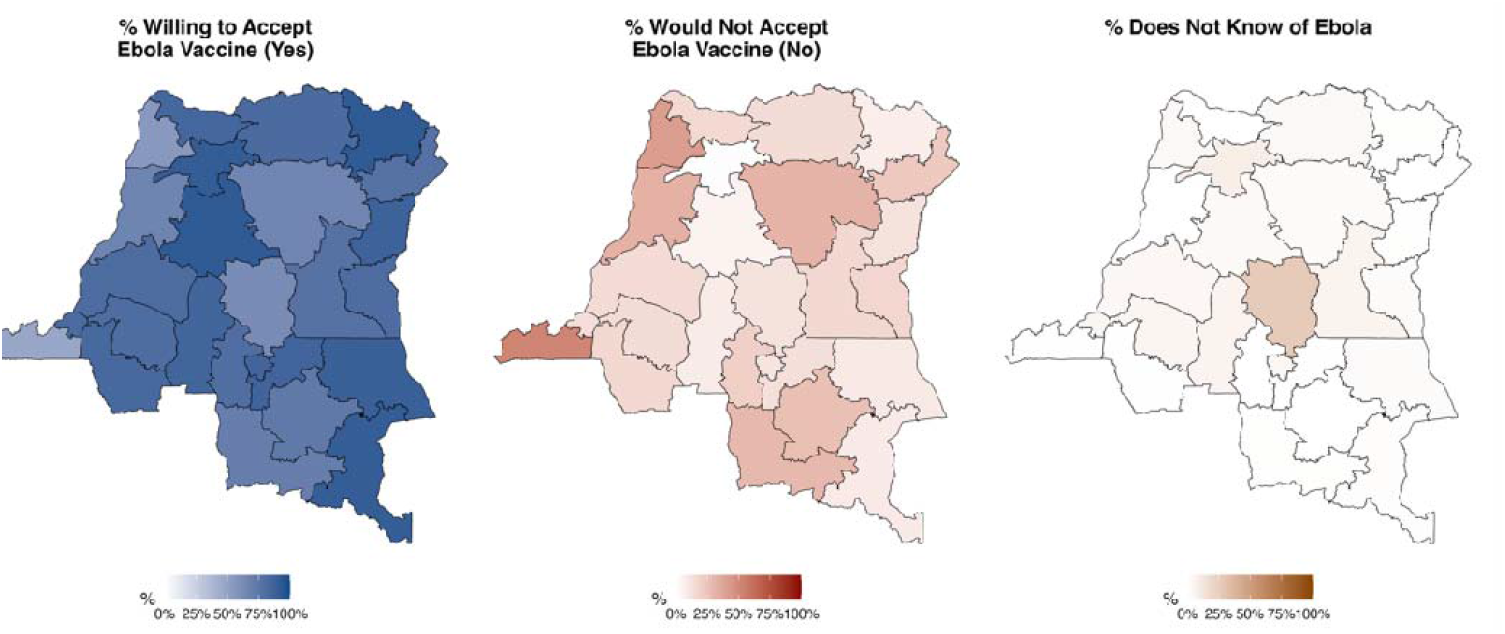
Geography of overall Ebola vaccine acceptance perceptions from a telephone phone survey conducted during December 2023–February 2024, by province, Democratic Republic of the Congo. A) Reported acceptance collapsed from the following three responses: Yes, for all populations; Yes, for children only; and Yes, for adults only. B) Reported nonacceptance by the response to: No, not interested. C) Reported knowledge of Ebola disease.

Ebola vaccine acceptance varied significantly by province (p<0.05) (Figure 1A). At present, seven provinces in DRC have reported confirmed BDBV infections during the current outbreak: Ituri (5,792 cases), Nord-Kivu (1,350 cases), Haut-Uélé (292 cases), Tshopo (30 cases), Bas-Uélé (seven cases), Sud-Kivu (three cases), and Sud-Ubangi (one case). Acceptance of an Ebola vaccine among the most affected provinces -Ituri, Nord-Kivu, and Haut-Uélé - was >75%.

However, vaccine acceptance rates among the remaining four provinces were more variable, ranging from 55.2% to 82.1%. Overall, males and females had nearly equivalent vaccine acceptance rates at 79.3% and 77.9%, respectively (Figure 2B). When stratified by education, those who reported less than high school, some college, or a Bachelors or advanced degree reported >80% acceptance; respondents who identified as having graduated high school had the lowest acceptance rate (70.4%). Across reported respondent occupations, Ebola vaccine acceptance was highest among HCWs (85.0%; 95% CI 83.7% – 86.3%) with the lowest rates among religious leaders (60.7%) and unemployed/”not working” (62.9%). Rural respondents had significantly higher acceptance rates than urban respondents: 82.6% and 75.4%, respectively (p<0.05). Ebola vaccine acceptance was also significantly higher among respondents who reported receiving a COVID-19 vaccine as compared to those who reported no COVID-19 vaccination: 81.7% and 61.2%, respectively. Lastly, Ebola vaccine acceptance was also higher among respondents who did not report an underlying health condition as compared to respondents with a health condition (80.2% vs. 68.4%) (Figure 1B).

## CONCLUSIONS

The ongoing Bundibugyo virus disease outbreak in the DRC is the largest Ebola disease outbreak in the country’s history, and the second largest ever recorded globally. The ERVEBO vaccine, which targets EBOV glycoprotein (GP) has been deployed during prior EVD outbreaks in theDRC. While there is limited information on the cross-protective activity against BDBV, 70,000 vaccine doses have been allocated to DRC, which include 20,000 for the Bundibugyo, Répondre Avec la Vaccination (BRAVO) Phase 3 clinical trial in frontline healthcare workers, and 50,000 for HCWs(14, 15). Given the heightened interest in broader Ebola vaccine deployment to help combat the ongoing outbreak, the expanding geography of the outbreak to include numerous provinces, and importance of community-centered approaches to increase vaccine uptake, we assessed Ebola vaccine trust and acceptance in the DRC. These observations build on our prior mpox vaccine trust and acceptance activities (17) to establish whole-of-country Ebola vaccine data across each province in the DRC.

Our data showed that vaccine acceptance among the most affected provinces in the current BVD outbreak, Ituri, Nord-Kivu, and Haut-Uélé was >75%, while vaccine acceptance rates were variable among the other affected provinces. Thus, these data suggest that future Ebola vaccine acceptance and uptake campaigns should consider provincial-level variabilities in their deployment strategies. While Ebola vaccine acceptance did vary significantly between rural and urban geographies, acceptance was >75% in both populations suggesting that vaccine acceptance and uptake variability between the provinces is not purely a function of greater rural or urban populations. Lastly, while our assessment of Ebola knowledge suggested that most provinces had strong knowledge of the disease, Sankuru was the lowest among all provinces, as was also observed in our prior assessment of mpox knowledge. While BVD has not yet been reported in Sankuru, it should be appreciated that lower disease knowledge as compared to other provinces, including those with confirmed infections, highlight the need to ensure disease-specific information is available in this region.

Limitations in this study included the requirement for participants to have telephone access, which may have reduced the inclusion of low socioeconomic status individuals, and an overrepresentation of men due to the focus of study recruitment on HCWs. However, given the focus of the initial Ebola vaccine deployments on this group, these data are likely highly pertinent. Overall, our data highlight that while Ebola vaccine acceptance and disease knowledge are relatively strong nationally in the DRC, including among the most affected provinces to date in the current BVD outbreak, ongoing geographic expansion could result in the need for enhanced community engagement activities.

## Data Availability

The data underlying these findings will be provided upon reasonable request to the corresponding author.

## ACKNOWLEDGEMENTS

We thank Brooke Aksnes and Emma Wray Aberle-Grasse for their continued support of this ongoing research endeavor. Moreover, this work would not have been possible without the tireless commitment of our interviewing staff, and dedicated time of all phone interview participants.

Ethics approval was provided by the Kinshasa School of Public Health (approval no. ESP/CE/188/2023) and the University of California, Los Angeles, as a public health response activity designation.

This work was supported by funding from the Centers for Disease Control and Prevention through the Task Force for Global Health (TFGH) under cooperative agreement no.

6NU2RGH001916-02-06. The content of the information does not necessarily reflect the position or the policy of TFGH, and no official endorsement should be inferred.

## BIOGRAPHY

Ms. Merritt is a PhD student in Epidemiology at the UCLA Fielding School of Public Health where her work examines the epidemiology, surveillance, and prevention of infectious diseases, with a particular focus in the Democratic Republic of the Congo. Dr. Kindrachuk is an associate professor and Canada Research Chair at the University of Manitoba, Winnipeg, Manitoba, Canada; his research interests focus on the investigation of emerging and re-emerging viruses.

